# Does prosperity pay? Unraveling the relationship between economic performance and life expectancy across a large number of European regions, 2008-2019

**DOI:** 10.1101/2024.06.11.24308750

**Authors:** Markus Sauerberg, Laura Ann Cilek, Michael Mühlichen, Florian Bonnet, Ina Alliger, Carlo Giovanni Camarda

**Affiliations:** Federal Institute for Population Research (BiB), Wiesbaden, Germany; French Institute for Demographic Studies (INED), Aubervilliers, France

**Keywords:** Life expectancy, gross domestic product, Preston curve, regional disparities, regression models, Europe

## Abstract

Understanding the relationship between life expectancy at birth (*e*_0_) and the gross domestic product per capita (GDPpc) is relevant for cohesion policies in the European Union (EU), because it might imply that economic convergence (or divergence) is accompanied by narrowing (or widening) health gaps. Previous studies have studied the association between GDPpc and *e*_0_ almost exclusively based on national data. However, it is certainly relevant to add a subnational dimension, because levels and trends in both *e*_0_ and GDPpc vary substantially across Europe’s regions. Accordingly, the aim of our study is examining whether the economic performance of a region is correlated to their *e*_0_ level. To do so, we collected official mortality and population counts from national statistical offices and information on GDPpc from the Eurostat database for 506 regions in 21 European countries from 2008 to 2019. Using this data, we built Preston curves from regression models. Our results suggest that there is indeed a positive association between GDPpc and *e*_0_. Similarly to Preston’s original analysis, we observe an upward shift in the curve, indicating that factors exogenous to a region’s GDPpc level also play an important role in explaining *e*_0_ gains. Yet, the relationship differs between geographical areas, and we also find examples, such as women in Germany, Austria, Poland, and the Netherlands, where the relationship/pattern does not seem to hold.

## 1. Introduction

About 50 years ago, Samuel H. Preston published his classic analysis on the association between mortality and level of economic development (Preston 1975). In his study, he deduces two key findings. First, there is a strong positive relationship between national GDPpc levels measured through GDP per capita (GDPpc) and life expectancy at birth (*e*_0_), which holds especially among low-GDPpc countries but becomes less pronounced as GDPpc rises. Second, he observes an upward shift in the relationship between a country’s GDPpc level and its *e*_0_ value over time. The change in the GDPpc-life expectancy relationship indicates that even those countries with no or very little economic improvements between two points in time have experienced mortality reductions. Preston therefore argues, that “factors exogenous to a nation’s level of GDPpc per head have had a major effect on mortality trends in more developed as well as in less developed countries” (Preston 1975: 243).

Over the last five decades, Preston’s analysis has sparked a debate on whether prosperity leads to higher levels of population health (Bloom and Canning 2007; Kunitz 2007; Mackenbach 2007). While McKeown (1976) has ascribed the success in reducing mortality almost entirely to improvements in living standards, Lutz and Kebede (2018) emphasize the role of educational expansion for progress in health and life expectancy. Furthermore, Wilkinson (1992) and Rogers (2002) have shown that GDPpc inequality (e.g., the Gini coefficient) is also an important predictor for a country’s mortality level and should be included in addition to GDPpc. By considering the GDPpc distribution of a given country, the magnitude of inequalities can be taken into account. For instance, a country might be comparatively wealthy measured through national GDPpc but its wealth is not equally distributed and mostly reserved for a privileged group. This country is likely to show a lower *e*_0_ value than suggested by its GDPpc level (Shkolnikov et al. 2019).

In the European context, Mackenbach and Looman (2013) have investigated to what extent the remarkable mortality reductions throughout the 20^th^ century had been accompanied by an upward shift in the relationship between GDPpc and *e*_0_. They found that improvements in longevity were independent of economic growth up to the 1960s. During the second half of the 20^th^ century, however, their empirical analysis suggests that gains in *e*_0_ were primarily driven by rises in national GDPpc. This makes sense given that Europe’s mortality pattern is characterized by a strong East-West gap with lower GDPpc levels and higher death rates found in former communist countries (Meslé and Vallin 2017; Grigoriev and Pechholdová 2017; Leon 2011). Those countries with rapid economic growth could reduce their death rates through improvements in living standards and access to medical innovations.

In more recent years, the relationship between GDPpc and *e*_0_ is less clear. As an example, countries with strong economies such as Germany, the UK or the US have been underperforming in terms of their *e*_0_ trends (Jasilionis et al. 2023; Grigoriev et al. 2024; Ho 2022). Yet, mortality variation within a country can be linked to the spatial distribution of social deprivation. This might indicate that relative GDPpc differences associated with social status are more important for explaining health disparities in developed societies (Wilkinson 1997). Accordingly, the aim of our paper is to examine the relationship between GDPpc and *e*_0_ from a regional perspective. Instead of focusing solely on national aggregates, we use GDPpc and mortality data for 506 spatial units in 21 European countries, covering the period from 2008 to 2019. The time span is convenient for studying the GDPpc-life expectancy relationship because it starts shortly after the EU enlargement to the east when disadvantaged regions received substantial support for economic growth within the EU’s cohesion policy plan. We do not include more recent years in our analysis so that our results are not affected by the COVID-19 pandemic.

## 2. Data and methods

Our empirical analysis is based on death and mid-year population counts by age and sex for 506 regions in 21 European countries (Austria, Belgium, Czech Republic, Denmark, Estonia, Finland, France, Germany, Hungary, Italy, Latvia, Lithuania, Netherlands, Norway, Poland, Portugal, Sweden, Slovenia, Slovakia, and Switzerland), which we obtained from the national statistical offices of these countries. Regions are defined in accordance with the EU’s Nomenclature of Territorial Units for Statistics (NUTS). Depending on the size of the country, we rely either on NUTS-2 or NUTS-3 levels. For Germany, however, we used a national classification developed for spatial planning purposes called ‘Raumordnungsregionen’, which is very similar in its size and structure to the NUTS-3 level in France and Spain. Each ‘Raumordnungsregion’ can be derived through aggregating the corresponding data at the NUTS-3 level. For Italy, we aggregated some NUTS-3 regions to avoid a break in the time series due to territorial changes. Moreover, due to data limitations, we cannot consider within-country variation for Estonia and Latvia in our analysis. Nevertheless, we consider both countries at the national level (NUTS-0) as they joined the EU in 2004, making their development in terms of GDPpc and *e*_0_ highly relevant for our study. More details about the regional division of Europe can be found in the appendix (Table A1).

To avoid annual fluctuations in the regional mortality data, we applied the smoothing method proposed by Camarda (2019). The model enables us to smooth the sex-specific death rates over age and time for all regions under study independently. Period life tables by sex, year, and region as well as the corresponding *e*_0_ estimates are derived on the basis of smoothed the death rates. Thus, our *e*_0_ figures do not reflect fluctuations in period mortality such as the *e*_0_ drop in 2015 for many European countries but rather correspond to the general trend of regional *e*_0_ over time. The national *e*_0_ value is given by the population-weighted average of regional *e*_0_ values. Aggregating the region-specific death and mid-year population counts to the national level and then applying the aforementioned smoothing method leads to very similar national *e*_0_ values. Yet, deriving national *e*_0_ from its population-weighted *e*_0_ values at the subnational level ensures consistency from a formal demography perspective (Vaupel and Canudas-Romo 2002).

The GDPpc data for different NUTS levels can be freely downloaded from the Eurostat database.

Finally, the relationship between GDPpc and *e*_0_ is studied on the basis of linear regression models,

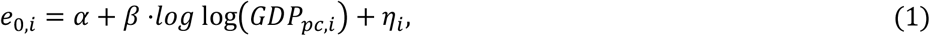

where *e*_0_,*i* represents the period life expectancy at birth estimate for region *i*, while *log(GDPpc*,*i)* refers to the corresponding log-transformed GDP per capita figure.

We fitted the regression model to both country-specific and regional data. To analyze east-west differences in the GDPpc-life expectancy relationship, we split our 506 regions into 389 regions located in the west (regions in Austria, Belgium, Denmark, Finland, France, Germany (West), Italy, Netherlands, Norway, Portugal, Sweden, and Switzerland) and 117 regions located in the east (Czech Republic, Estonia, Germany (East), Hungary, Latvia, Lithuania, Poland, Slovenia, and Slovakia). In addition, we fitted a fixed-effect panel regression model to study the relationship between GDPpc and *e*_0_ in a longitudinal setting. The model specification corresponds to equation (1). All statistical analyses presented in this paper have been conducted in R (R Core Team 2024).

## 3. Results

The national *e*_0_ value is positively associated with a country’s GDPpc level in all analyzed years. The curves drawn in Figure 1 represent the fitted regression lines to cross-sectional data for the years 2008 to 2019. The dots correspond to observed data and the color indicate the year of observation. For instance, black-colored dots correspond to data observed in the year 2008, while dots colored in light yellow indicate that the data refers to the year 2019. The relationship between *e*_0_ and GDPpc holds for both women and men but is stronger among men. The curves drawn in the figure suggest an upwards shift in the relationship over time. The corresponding *β* coefficients and *p* values are presented in the appendix. For men, the *β* coefficients range from 4.8 (in 2013) to 5.7 (in 2008), and the model can explain about 67 to 77 percent of the variation in the data, measured through *R*^*2*^. For instance, the model fitted to data observed in 2013 suggests that a 10 percent increase in GDPpc corresponds to an expected *e*_0_ value by about 0.5 years.

**Figure 1:**
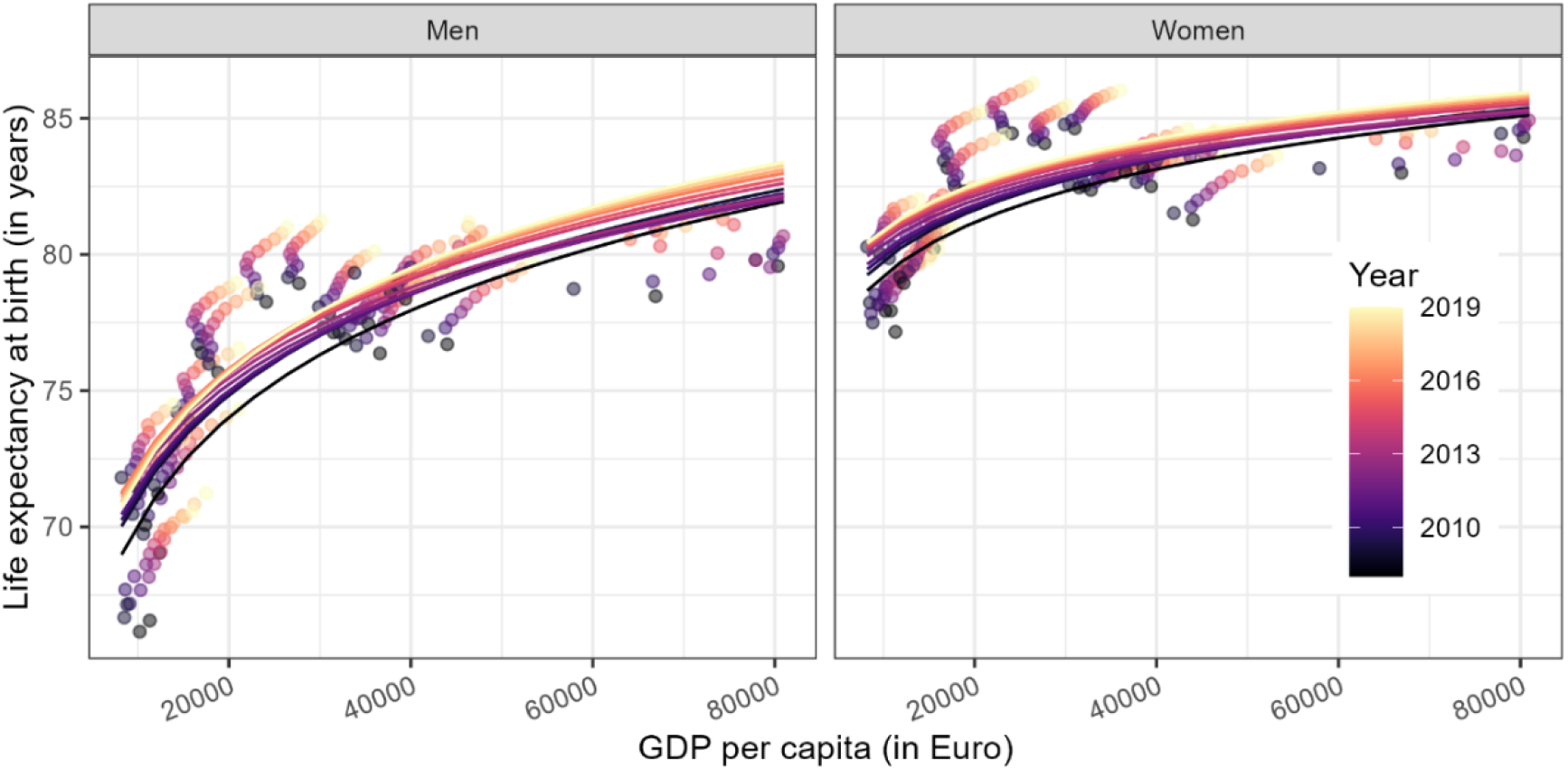
Period life expectancy at birth (*e*_*0*_) regressed on GDP per capita for 21 European countries, 2008 to 2019. Source: Mortality data comes from statistical offices (own calculations) and GDP per capita data was obtained from Eurostat (2024).

Among women, the estimated slope parameters as well as *R*^*2*^ are both slightly lower than for men, i.e., the *β* coefficient ranges from 2.3 (in 2014) to 2.8 (in 2008), while *R*^*2*^ lies between 43 and 62 percent. Using regional data instead of national GDPpc and *e*_0_figures increases the number of observations substantially from 21 countries to 506 regions (see Figure 2). For both women and men, the *β* coefficients slightly decrease compared to the models fitted with country-level data. We still observe a strong positive relationship between GDPpc and *e*_0_ for both sexes (see table 1).

**Figure 2:**
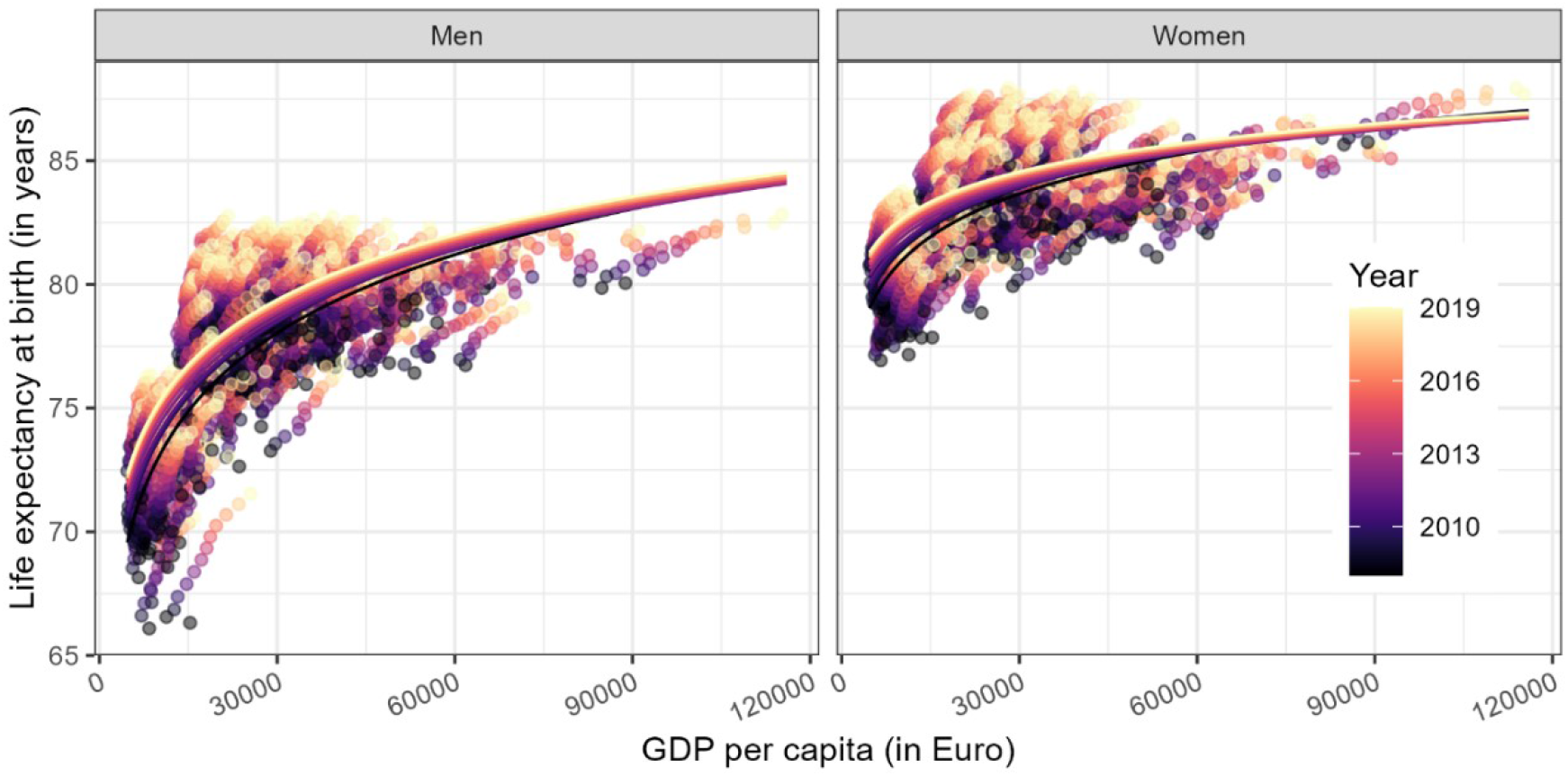
Period life expectancy at birth (*e*_*0*_) regressed on GDP per capita for 506 European regions, 2008 to 2019. Source: As of Figure 1.

**Figure 3:**
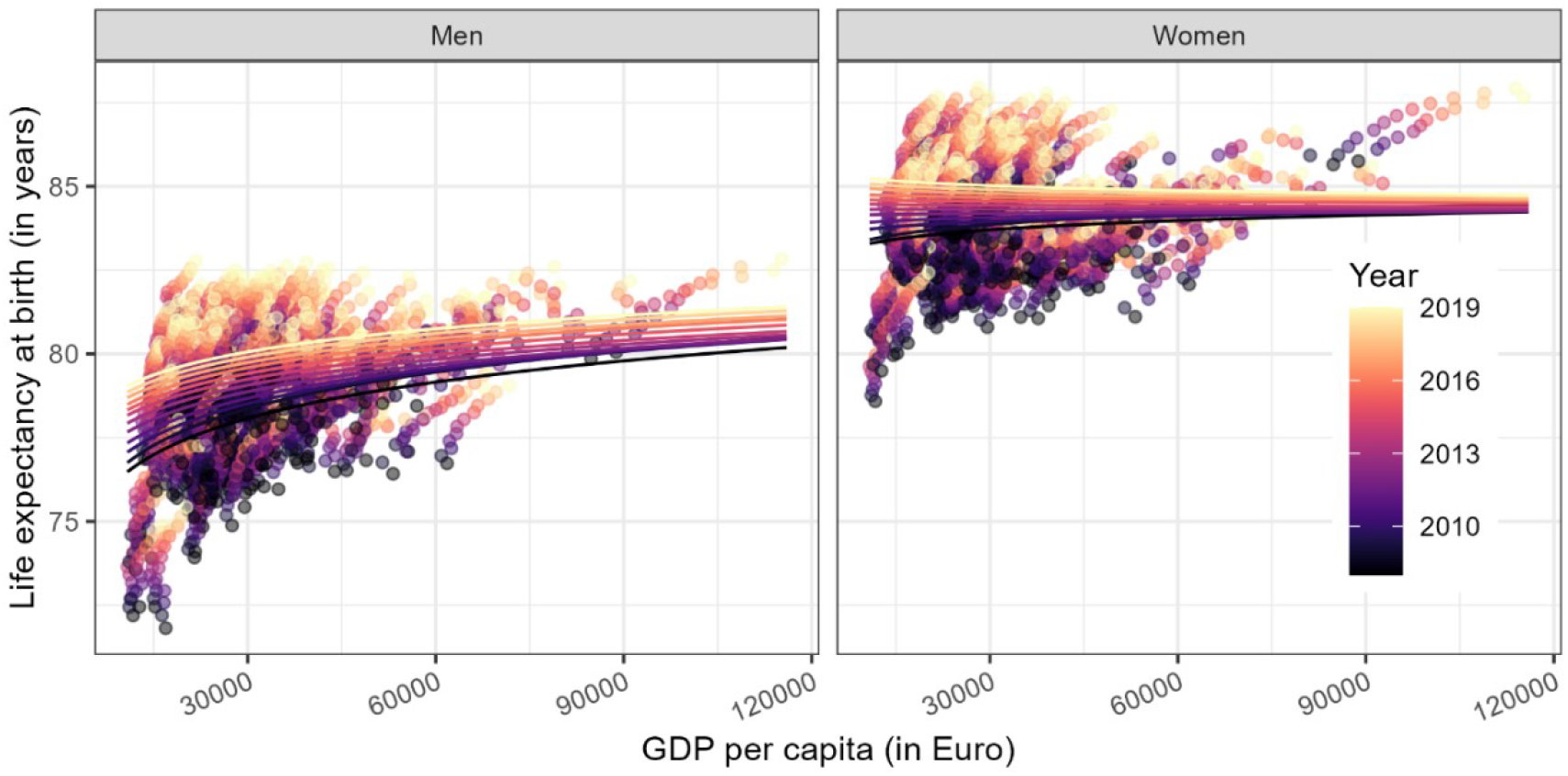
Period life expectancy at birth (*e*_*0*_) regressed on GDP per capita for 389 western European regions, 2008 to 2019. Source: As of Figure 1.

**Figure 4:**
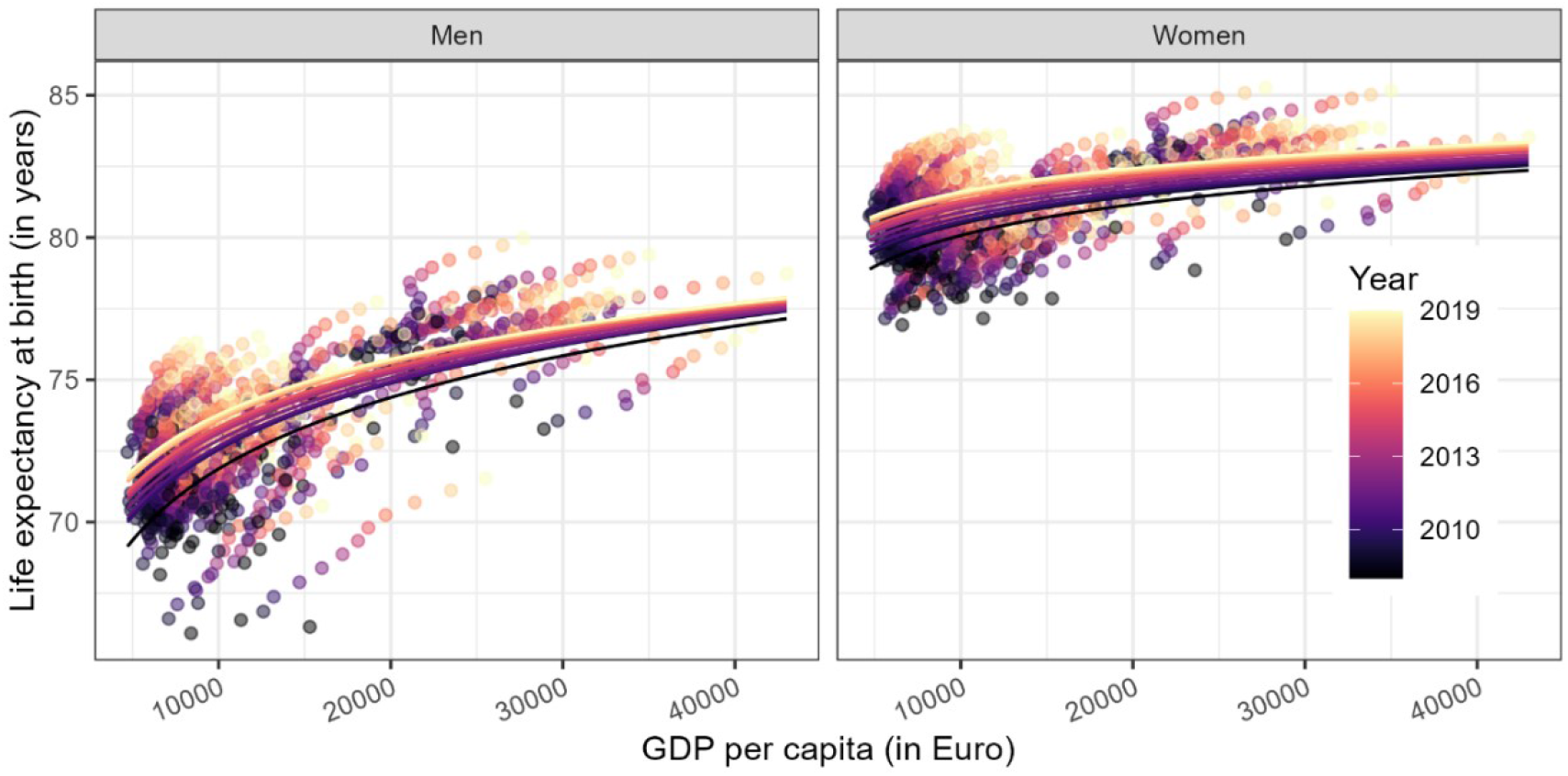
Period life expectancy at birth (*e*_*0*_) regressed on GDP per capita for 117 eastern European regions, 2008 to 2019. Source: As of Figure 1.

After splitting the data into two groups, (a) regions in former-communist countries and (b) the remaining regions, the region-specific regression models indicate that the relationship between GDPpc and *e*_0_ is primarily visible for former-communist regions. This holds especially true for women. Both the estimated *β* coefficients and the *R*^*2*^ measure for models based on data for women in western regions are close to zero, suggesting no statistical association between GDPpc and mortality.

When analyzing the GDPpc-life expectancy relationship within individual countries, we observe large cross-country differences. For instance, the relationship is comparatively strong in Belgium, Hungary, France, Germany, and Czechia, but less pronounced in the Netherlands, Poland, and Austria (see figure 5). Please note, however, that the number of regions within a given country varies considerably, i.e., the fitted regression models are based on a different number of observations. Detailed statistics for each fitted regression model can be found in the appendix (Table A2 and A3).

**Figure 5:**
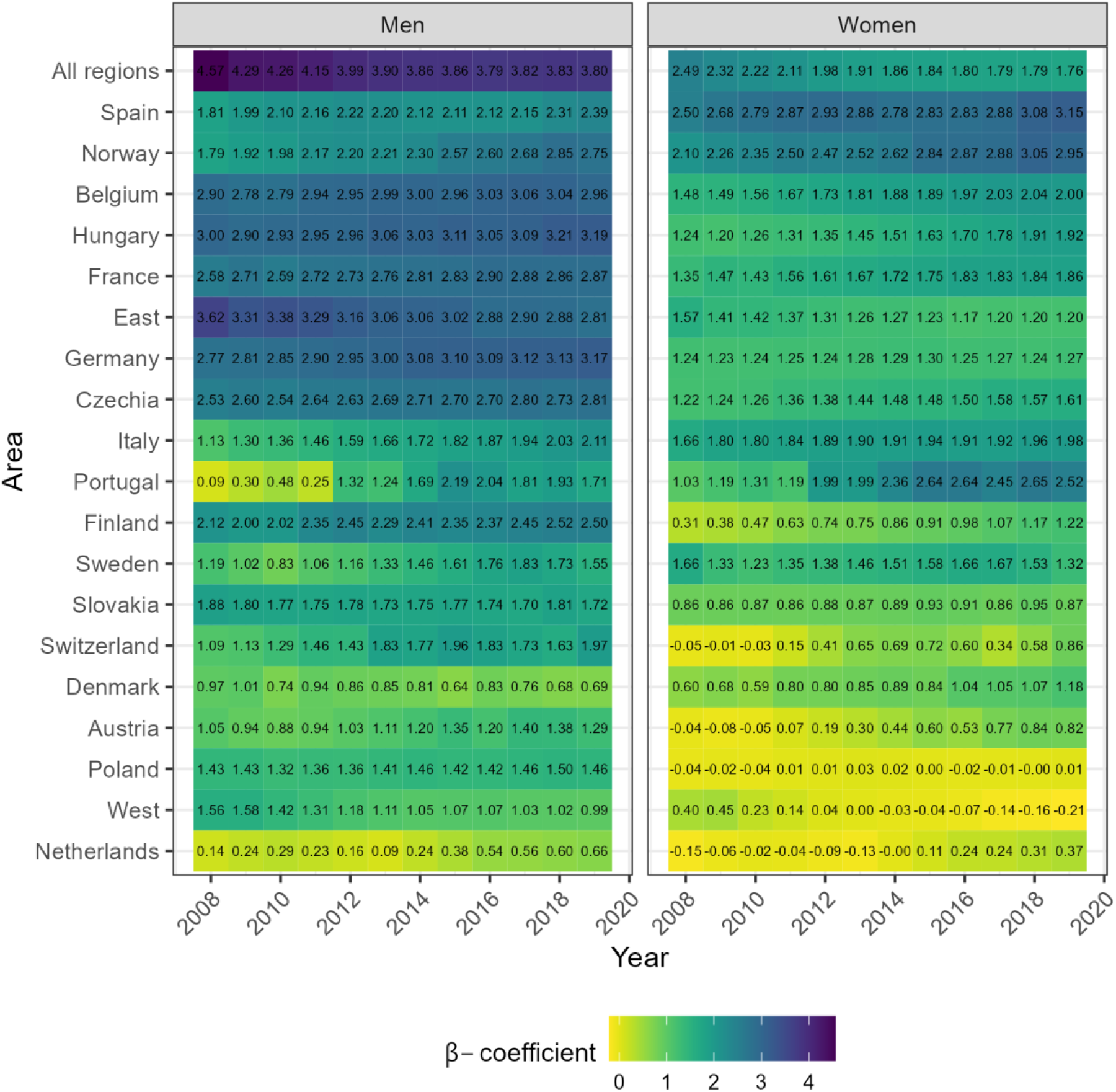
The *β* relationship between GDP per capita (GDPpc) and life expectancy at birth (*e*_*0*_) based on regional data in individual countries, 2008 to 2019. Source: As of Figure 1.

Interestingly, Figure 5 suggests an increase in the relationship over time for several countries. This can be observed, for example, in Germany, Belgium, and Hungary among men and in Spain, Norway, and Portugal among women. However, when considering all regions, the β coefficient decreases over time for both men and women, highlighting distinct patterns in the relationship depending on the population study. Finally, we studied the correlation between GDPpc and *e*_0_ in a longitudinal setting. The regression results are presented in Figure 6, indicating that those countries with increases in their GDPpc level are also those countries that show gains in *e*_0_. Again, the relationship is stronger among men. We do not observe a clear east-west pattern. But, in contrast to the cross-sectional regression results, our panel regression results suggest rather high associations between GDPpc and *e*_0_ for western and Nordic European countries such as France, Finland, and Denmark. Nevertheless, the results from the country-specific models vary considerably, with the highest *β* coefficient found for men in Denmark (11.7) and the lowest found for women in Switzerland (2.7). In other words, data for Danish men suggests that a one percent increase in GDPpc corresponds to a*e*_0_ gain of about 0.1 years, whereas the *e*_0_ increase using data for women in Switzerland is about 0.03 years (see table A4). Accordingly, the results depend on how much GDPpc and *e*_0_ have grown between 2008 and 2019 in the given country.

**Figure 6:**
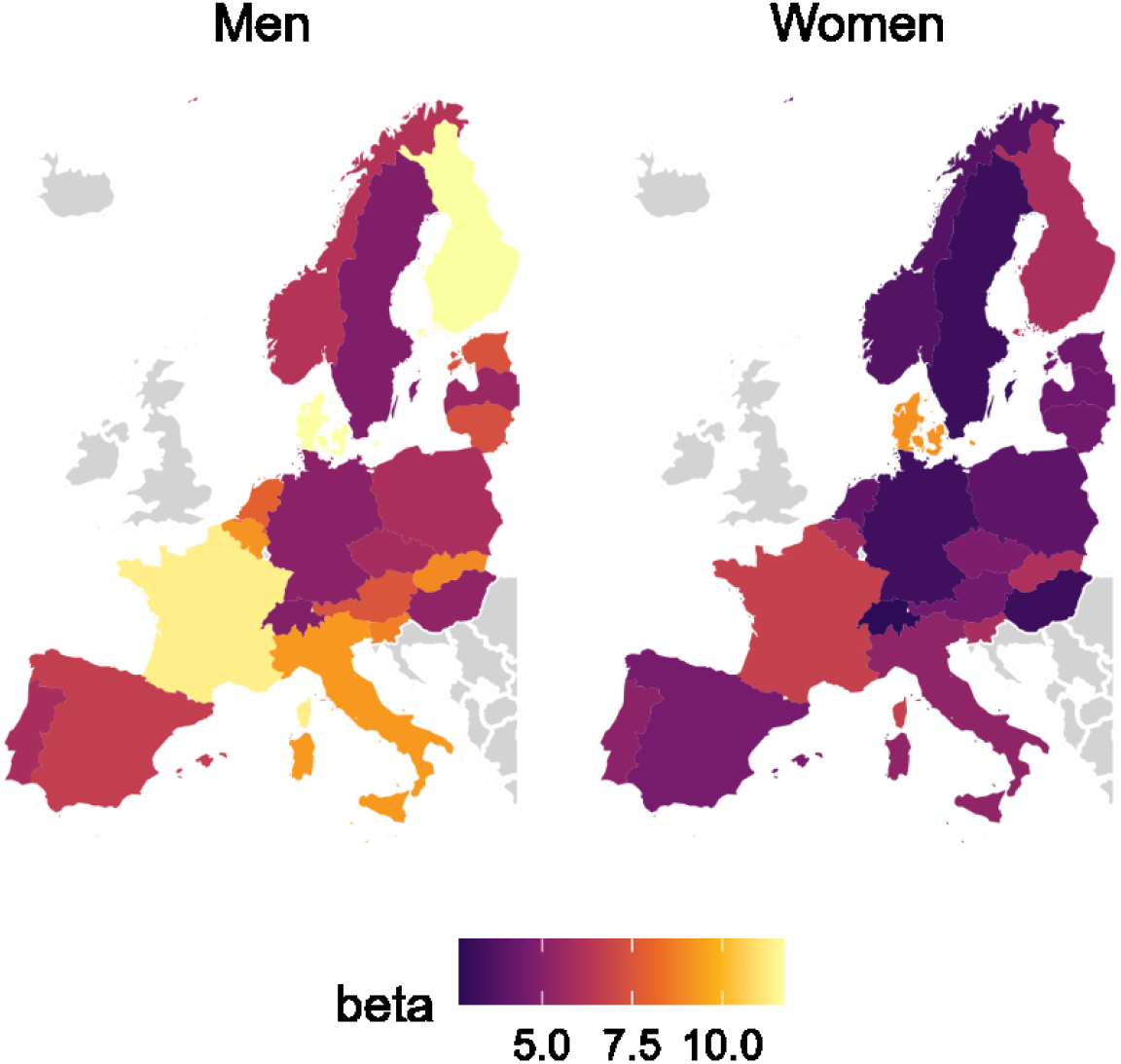
β Association between GDP per capita (GDPpc) and life expectancy at birth (*e*_*0*_) based on longitudinal data for 506 regions in 21 countries, 2008 to 2019. Source: As of Figure 1.

Finally, we present time trends in the regional variation of *e*_0_ and GDPpc in Figure 7 and 8. The graphs show the Gini coefficient with respect to *e*_0_ or GDPpc from 2008 to 2019 for all regions as well as for regions located split based on their location in western or eastern Europe. The level of regional variation in *e*_0_ is substantially larger for eastern regions compared to the regions in the west. Further, regional variation in *e*_0_ decreased between 2008 and 2019 mainly in the eastern part of Europe. For regions in the west, regional variation in *e*_0_ stagnated or even slightly increased over the analyzed time period.

**Figure 7:**
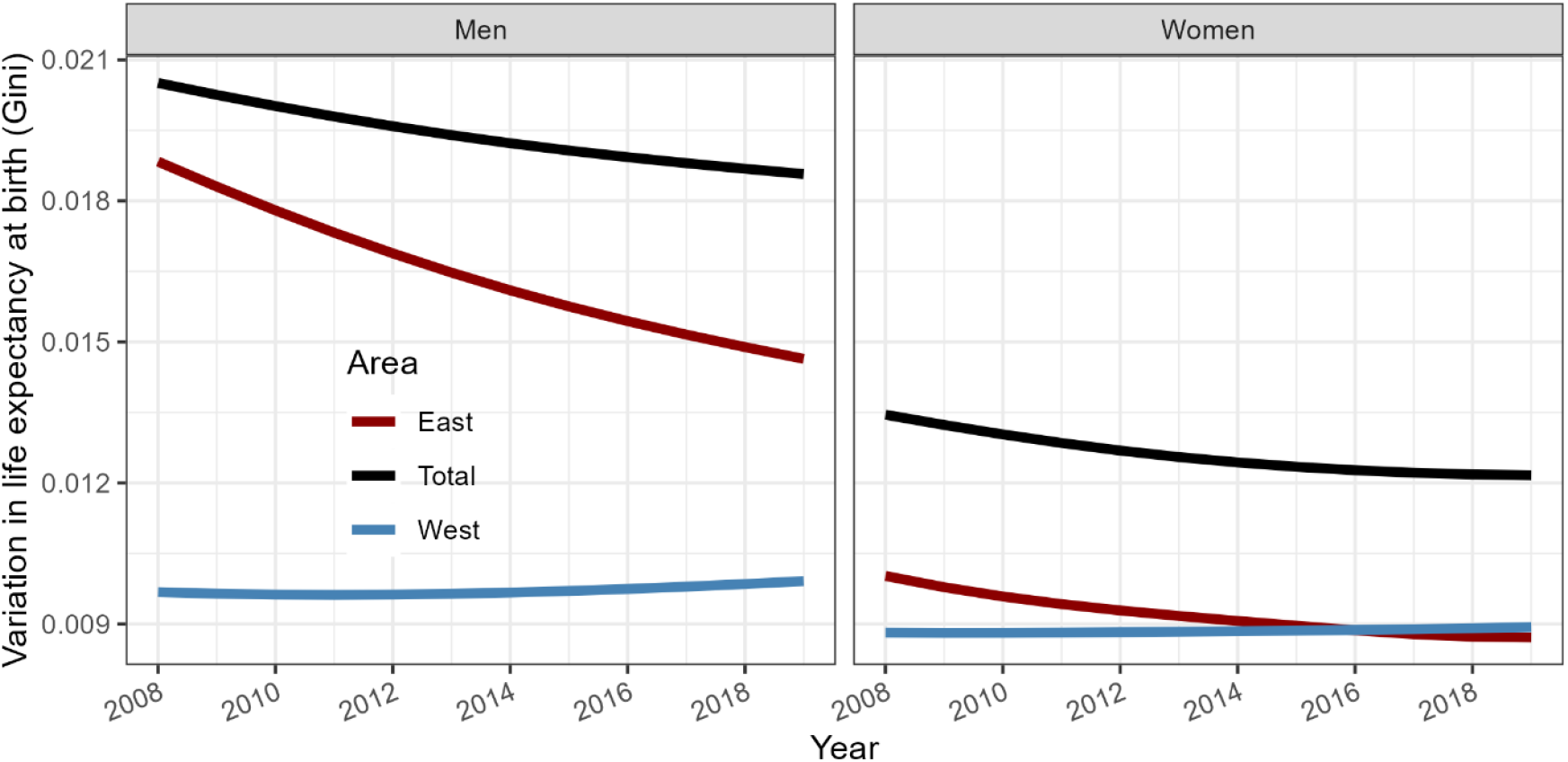
Regional variation in life expectancy at birth (*e*_*0*_) measured through the Gini coefficient, 2008 to 2019. Source: Mortality data comes from statistical offices (own calculations).

**Figure 8:**
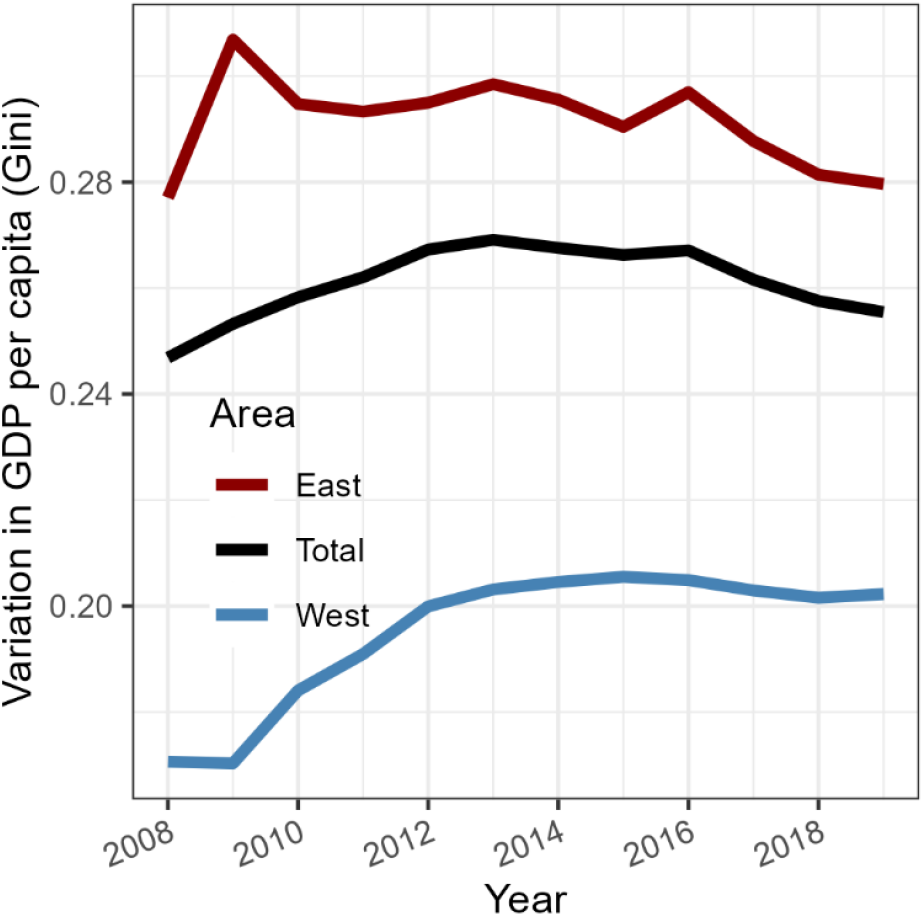
Regional variation in GDP per capita (GDPpc) measured through the Gini coefficient, 2008 to 2019. Source: Eurostat data (2024).

The regional variation in GDPpc is larger for eastern regions than for western regions (see Figure 8). The time trends reveal that regional variation in GDPpc increased during the earlier years of the analyzed time span. In more recent years, however, regional variation in GDPpc tends to decline in eastern regions and stagnates in western regions.

## 4. Discussion

### Summary of main findings

In our study, we examined the relationship between economic performance measured through GDPpc and *e*_0_ at the subnational level for 21 European countries. Linear regression models in a cross-sectional setting based on data for 506 spatial units reveal that there is a positive association between a region’s economic performance and its *e*_0_ level. In line with Preston’s analysis, we observe an upward shift in the relationship between a regions’s GDPpc level and its *e*_0_ value over time. The Preston-relationship holds for both sexes but appears to be more pronounced among men.

After splitting the 506 spatial units into two groups, (a) regions located in those countries which joined the EU in 2004 and (b) regions located in those countries which were already EU member states before 2004, then performing the same analysis, we find that the GDPpc-life expectancy relationship remains observable for newer EU member states but is very weak or even non-existing (among women) in older EU member states. Looking at the correlation between a regions’s GDPpc level and its *e*_0_ value within specific countries further reveals that there is considerable variability within each of the two groups. For instance, the GDPpc-life expectancy relationship is comparatively weak for regions in Poland – although joined the EU in 2004, whereas the relationship is rather strong in France.

Finally, we studied the association between GDPpc and *e*_0_ value in a longitudinal setting, i.e., applying fixed effects panel regressions. Again, we find empirical evidence for positive correlation between the two variables for both women and men. The magnitude of the correlation differs substantially across the 21 analyzed countries, with highest *β* coefficients found for France, Finland, and Denmark, while *β* coefficients are comparatively low for panel regression models based on Swedish and Swiss data. Accordingly, we do not identify a clear geographical pattern for our panel regression results.

### Interpretation

Our results are partly in line with Preston’s original analysis. Especially among lower-GDPpc countries, we observe a clear relationship between GDPpc and *e*_0_. Indeed, we observe an upward shift in the relationship between a country’s GDPpc level and its *e*_0_ value over time, suggesting that gains in *e*_0_ cannot be ascribed to rising GDPpc levels alone. Some possible explanations are the health measures implemented in central-eastern European countries after joining the EU in 2004, e.g., enhancements in road safety, adoption of alcohol-control policies, and general improvements in access to a modern healthcare system (Trias Llimos et al 2018; Hrzic 2021; Lai and Habicht 2011; Jasilionis, Leon, and Pechholdová 2020). Yet, it is also important to consider that many European countries were facing a financial crisis in 2007 and 2008, which led to large drops in GDP levels for many European regions (Crescenzi, Luca, and Milio 2016). It is likely that this has contributed to the upward shift in the GDPpc-life expectancy relationship.

Further, we find countries and regions where the GDPpc-life expectancy relationship is comparatively weak or does not hold at all, such as among women in Germany and Austria. The eastern part of Germany is economically disadvantaged compared to the western part of the country. Among men, the east-west divide is also visible in terms of mortality, which makes GDPpc a good predictor for a region’s *e*_0_ level for men in Germany. For women, however, mortality levels are not higher in eastern Germany than in the western part. In some regions, mortality is even lower in the east as compared to the west. The observed geographical pattern in mortality has been attributed to differences in the past smoking history of women in eastern and western Germany (Vogt et al. 2017). Accordingly, the GDPpc-life expectancy relationship does not hold for women in Germany because social norms, lifestyle choices, and health behavior offset the (dis)advantages in a region’s GDPpc level. In Austria, we observe a particularly large GDPpc level for the capital region of Vienna. Interestingly, mortality is much higher in Vienna compared to other regions in Austria, leading to a rather weak GDPpc-life expectancy relationship. This might change by taking into account the within-region variability in both GDPpc and mortality in Vienna. In other words, it is likely that those districts in Vienna with an advantage in their socioeconomic situation are have above-average *e*_0_ levels.

Moreover, we observe sex differences in the GDPpc-life expectancy relationship. Generally speaking, the relationship is stronger among men. This may be attributed to higher variability in mortality among men (Mackenbach et al. 1999; Sauerberg et al. 2023). Mortality rates are particularly elevated for those men with a low socioeconomic status, thus reinforcing the positive correlation between a region’s GDPpc and *e*_0_ level (Luy and Gast 2014; Anson 2003). Still, we find that in some countries, such as Spain and Norway, the GDPpc-life expectancy relationship is stronger among women than men. It might be worthwhile investigating whether this can be explained through higher employment rates for women in these countries.

### Strengths and limitations

Our analysis is based on high-quality data for a large number of European regions, including all countries which joined the EU in 2004. The analyzed time span, 2008 to 2019, is interesting because it can help identify potential effects of the EU enlargement. Nonetheless, both local and international economic shocks are not uncommon in today’s global economy, so it may be worth additionally exploring the relationship between *e*_0_ and GDPpc in countries during and after periods of economic turmoil such as the financial crisis in 2008. For example, it may be useful to understand if *e*_0_ in regions with high GDPpc is more (or less) resilient to economic shocks and subsequent recovery than in areas with lower GDPpc. In addition, the number of regions in some analyzed countries such as Denmark or Finland is relatively low. Statistical results based on a low number of observations should not be overinterpreted. While our regression models do not control for spatial effects, robustness checks indicate that the direction of the relationship remains the same when spatial autocorrelation is controlled for (not shown). Finally, it should be noted that we have not yet considered any GDPpc inequality measures such as the Gini coefficient, even though previous research has demonstrated its relevance in examining the GDPpc-life expectancy relationship (Torre and Myrskylä 2024; Wilkinson 1992; Rogers 2002). The reason for that is limited data on the Gini coefficient at the subnational level. Moreover, our analysis cannot claim any causality.

## 5. Conclusions

About 50 years after Preston’s influential analysis, the relationship between GDPpc and *e*_0_ still holds for Europe’s regions. However, in specific subsets of our data, such as women in Germany, Austria, Poland, and the Netherlands, we find that higher GDPpc levels are not necessarily associated with higher *e*_0_ values. This indicates that the relationship is not fixed but rather varies between countries and across time. The fact that we do not observe the GDPpc-life expectancy relationship for women in the west of Europe may suggest that GDPpc might become less relevant as a predictor for *e*_0_ in populations with lowest-low mortality levels.

## Data Availability

All data produced in the present study are available upon reasonable request to the authors.

## Acknowledgements

We are very grateful to Markéta Majerová, Rok Hrzic, Magdalena Muszyńska-Spielauer, and Mathias Lerch for providing, respectively, Czech, Slovenian, Austrian, and Swiss data. I.A., M.M., and M.S. were supported by funding from the European Research Council (ERC) under the European Union’s Horizon 2020 research and innovation programme (grant agreement No 851485).

## Appendix

**A1:** Regional classification for 21 European countries.

A1: Regional classification for 21 European countries
| Country | NUTS level | “East” or “West” | Number of spatial units | Adjustments or comments |
| --- | --- | --- | --- | --- |
| Austria | 2 | West | 9 |  |
| Belgium | 2 | West | 11 |  |
| Switzerland | 2 | West | 7 |  |
| Czechia | 2 | East | 8 |  |
| Germany (Total) | ROR |  | 96 | Harmonized to apply current territorial administrative division (as of Dec 2022) to the whole study period and to eliminate the Census 2011 break; 400 NUTS 3 units (‘Kreise’) aggregated to 96 ROR units according to classification of BBSR (2017). |
| Germany (East) | ROR | East | 22 |  |
| Germany (West) | ROR | West | 74 |  |
| Denmark | 2 | West | 5 |  |
| Estonia | 0 | East | 1 |  |
| Spain | 3 | West | 50 | For visibility reasons, we excluded the Canary Islands. |
| Finland | 2 | West | 4 | We merged Åland Islands to South Finland. |
| France | 3 | West | 95 | Non-European areas excluded; north and south of Corse merged to maintain consistent time series. |
| Hungary | 2 | East | 8 |  |
| Italy | 3 | West | 92 | We merged the following regions to maintain a consistent time series: 1) Biella + Vercelli, 2) Novara + Verbano, 3) Como + Lecco, 4) Milano + Lodi + Monza + Brianza, 5) Rimini + Forli-Cesena, 5) Firenze + Prato, 6) Cagliari + Medio Campidano + Carbonia-Iglesias + Ogliastra + Oristano + Nuoro, 7) Sassari + Olbia-Tempio, 8) Foggia + Bari + Barletta, 9) Fermo + Ascoli-Piceno, 10) Crotone + Vibo Valentia + Cantanzaro. |
| Lithuania | 2 | East | 2 |  |
| Latvia | 1 | East | 1 |  |
| Netherlands | 2 | West | 12 |  |
| Norway | 2 | West | 7 | Harmonized to apply current territorial administrative divisions; we excluded remote islands Svalbard and Jan Mayen. |
| Poland | 3 | East | 73 |  |
| Portugal | 2 | West | 7 | For visibility reasons, we excluded the remote islands Azores and Madeira in our mortality maps. |
| Sweden | 2 | West | 8 |  |
| Slovenia | 2 | East | 2 |  |
| Slovakia | 3 | East | 8 |  |
| *Total N* |  |  | *506* |  |

**Table A2:** Cross-sectional linear regression statistics for 21 European countries, 2008 to 2019.

| Year | Men |  |  | Women |  |  |
| --- | --- | --- | --- | --- | --- | --- |
|  | slope | *p* value | *R*<sup>2</sup> | slope | *p* value | *R*<sup>2</sup> |
| 2008 | 5.66018 | 0.00000 | 0.72177 | 2.81421 | 0.00005 | 0.58989 |
| 2009 | 5.40565 | 0.00000 | 0.77264 | 2.67729 | 0.00002 | 0.62412 |
| 2010 | 5.23761 | 0.00000 | 0.77024 | 2.54639 | 0.00004 | 0.59672 |
| 2011 | 5.10249 | 0.00000 | 0.73837 | 2.45166 | 0.00010 | 0.55631 |
| 2012 | 4.85954 | 0.00000 | 0.69077 | 2.30097 | 0.00032 | 0.50351 |
| 2013 | 4.78779 | 0.00000 | 0.67956 | 2.24955 | 0.00044 | 0.48672 |
| 2014 | 4.98802 | 0.00000 | 0.68263 | 2.31747 | 0.00052 | 0.47821 |
| 2015 | 5.05702 | 0.00000 | 0.69130 | 2.32596 | 0.00053 | 0.47650 |
| 2016 | 5.11992 | 0.00000 | 0.69583 | 2.33792 | 0.00054 | 0.47634 |
| 2017 | 5.25593 | 0.00000 | 0.68698 | 2.36159 | 0.00073 | 0.45994 |
| 2018 | 5.40009 | 0.00000 | 0.67547 | 2.39414 | 0.00095 | 0.44519 |
| 2019 | 5.51557 | 0.00001 | 0.66578 | 2.41314 | 0.00121 | 0.43196 |

**Table A3:** Cross-sectional linear regression statistics for all regions, eastern regions, and western regions, 2008 to 2019.

| Year | Men |  |  | Women |  |  |
| --- | --- | --- | --- | --- | --- | --- |
|  | slope | *p* value | *R*<sup>2</sup> | slope | *p* value | *R*<sup>2</sup> |
| All regions |  |  |  |  |  |  |
| 2008 | 4.56540 | 0.00000 | 0.65094 | 2.48909 | 0.00000 | 0.43778 |
| 2009 | 4.29131 | 0.00000 | 0.67472 | 2.31947 | 0.00000 | 0.44981 |
| 2010 | 4.26080 | 0.00000 | 0.64051 | 2.21861 | 0.00000 | 0.39900 |
| 2011 | 4.14656 | 0.00000 | 0.61839 | 2.11260 | 0.00000 | 0.37057 |
| 2012 | 3.99444 | 0.00000 | 0.59075 | 1.98437 | 0.00000 | 0.33745 |
| 2013 | 3.89910 | 0.00000 | 0.57599 | 1.90891 | 0.00000 | 0.31960 |
| 2014 | 3.85881 | 0.00000 | 0.55646 | 1.86467 | 0.00000 | 0.30020 |
| 2015 | 3.85583 | 0.00000 | 0.54715 | 1.84468 | 0.00000 | 0.28829 |
| 2016 | 3.79185 | 0.00000 | 0.54933 | 1.80289 | 0.00000 | 0.28444 |
| 2017 | 3.82112 | 0.00000 | 0.52708 | 1.79407 | 0.00000 | 0.26431 |
| 2018 | 3.83356 | 0.00000 | 0.51170 | 1.79131 | 0.00000 | 0.25186 |
| 2019 | 3.79669 | 0.00000 | 0.49108 | 1.76444 | 0.00000 | 0.23630 |
| East |  |  |  |  |  |  |
| 2008 | 3.61768 | 0.00000 | 0.48848 | 1.57033 | 0.00000 | 0.27455 |
| 2009 | 3.31341 | 0.00000 | 0.52862 | 1.40956 | 0.00000 | 0.28334 |
| 2010 | 3.37622 | 0.00000 | 0.53316 | 1.42049 | 0.00000 | 0.27568 |
| 2011 | 3.29033 | 0.00000 | 0.52691 | 1.37363 | 0.00000 | 0.26292 |
| 2012 | 3.16025 | 0.00000 | 0.51734 | 1.30903 | 0.00000 | 0.24803 |
| 2013 | 3.06495 | 0.00000 | 0.51886 | 1.25911 | 0.00000 | 0.23839 |
| 2014 | 3.05896 | 0.00000 | 0.52766 | 1.26631 | 0.00000 | 0.23999 |
| 2015 | 3.02197 | 0.00000 | 0.51835 | 1.23440 | 0.00000 | 0.22434 |
| 2016 | 2.87967 | 0.00000 | 0.51078 | 1.17205 | 0.00000 | 0.21500 |
| 2017 | 2.90215 | 0.00000 | 0.50441 | 1.20207 | 0.00000 | 0.21542 |
| 2018 | 2.88442 | 0.00000 | 0.49348 | 1.19968 | 0.00000 | 0.20761 |
| 2019 | 2.81098 | 0.00000 | 0.47869 | 1.20293 | 0.00000 | 0.20700 |
| West |  |  |  |  |  |  |
| 2008 | 1.56235 | 0.00000 | 0.11598 | 0.40127 | 0.06685 | 0.00865 |
| 2009 | 1.57637 | 0.00000 | 0.11911 | 0.44539 | 0.04135 | 0.01071 |
| 2010 | 1.41963 | 0.00000 | 0.11010 | 0.23167 | 0.25897 | 0.00329 |
| 2011 | 1.30537 | 0.00000 | 0.10007 | 0.14436 | 0.46652 | 0.00137 |
| 2012 | 1.17820 | 0.00000 | 0.08879 | 0.04263 | 0.82256 | 0.00013 |
| 2013 | 1.11058 | 0.00000 | 0.08131 | 0.00381 | 0.98377 | 0.00000 |
| 2014 | 1.05428 | 0.00000 | 0.07460 | -0.03161 | 0.86463 | 0.00008 |
| 2015 | 1.06771 | 0.00000 | 0.07544 | -0.03701 | 0.84269 | 0.00010 |
| 2016 | 1.07188 | 0.00000 | 0.07466 | -0.06856 | 0.71512 | 0.00034 |
| 2017 | 1.02589 | 0.00000 | 0.06604 | -0.14434 | 0.44900 | 0.00148 |
| 2018 | 1.02137 | 0.00000 | 0.06361 | -0.16286 | 0.39857 | 0.00184 |
| 2019 | 0.99345 | 0.00000 | 0.05944 | -0.20623 | 0.28676 | 0.00293 |

**Table A4:** Longitudinal regression statistics based on data for all regions, eastern regions, western regions, and for individual countries, 2008 to 2019.

|  | Men |  |  | Women |  |  |
| --- | --- | --- | --- | --- | --- | --- |
|  | slope | *p* value | *R*<sup>2</sup> | slope | *p* value | *R*<sup>2</sup> |
| All regions | 6.44730 | 0.00000 | 0.60403 | 4.12515 | 0.00000 | 0.58847 |
| East | 5.97943 | 0.00000 | 0.79220 | 4.00341 | 0.00000 | 0.76174 |
| West | 6.94533 | 0.00000 | 0.51301 | 4.25475 | 0.00000 | 0.48531 |
| Austria | 7.48112 | 0.00000 | 0.93842 | 4.43042 | 0.00000 | 0.91264 |
| Belgium | 9.16791 | 0.00000 | 0.87750 | 5.69642 | 0.00000 | 0.83941 |
| Switzerland | 4.89798 | 0.00000 | 0.82225 | 2.72198 | 0.00000 | 0.79887 |
| Czechia | 5.81499 | 0.00000 | 0.67086 | 4.77645 | 0.00000 | 0.68322 |
| Germany | 5.12477 | 0.00000 | 0.90636 | 3.24585 | 0.00000 | 0.89204 |
| Denmark | 11.71662 | 0.00000 | 0.92361 | 9.07867 | 0.00000 | 0.91584 |
| Spain | 6.68450 | 0.00000 | 0.25906 | 4.61637 | 0.00000 | 0.25490 |
| Finland | 11.68991 | 0.00000 | 0.82362 | 5.97319 | 0.00000 | 0.78271 |
| France | 11.34632 | 0.00000 | 0.69505 | 6.74903 | 0.00000 | 0.67004 |
| Hungary | 5.26340 | 0.00000 | 0.63704 | 3.11847 | 0.00000 | 0.68378 |
| Italy | 9.29146 | 0.00000 | 0.34836 | 5.21793 | 0.00000 | 0.31322 |
| Netherlands | 7.90600 | 0.00000 | 0.58432 | 4.10284 | 0.00000 | 0.56274 |
| Norway | 6.24244 | 0.00000 | 0.39886 | 3.72584 | 0.00000 | 0.39504 |
| Poland | 5.98156 | 0.00000 | 0.80331 | 3.95661 | 0.00000 | 0.73798 |
| Portugal | 5.91068 | 0.00000 | 0.42573 | 5.13672 | 0.00000 | 0.51651 |
| Sweden | 4.82945 | 0.00000 | 0.56551 | 3.10394 | 0.00000 | 0.54986 |
| Slovakia | 8.89513 | 0.00000 | 0.90725 | 6.06525 | 0.00000 | 0.90538 |

## Notes

### Competing Interest Statement

The authors have declared no competing interest.

### Funding Statement

This study was supported by funding from the European Research Council (ERC) under the European Union's Horizon 2020 research and innovation programme (grant agreement No 851485).

### Author Declarations

National statistical offices and Eurostat database.

